# Severe SARS-CoV-2 infection in humans is defined by a shift in the serum lipidome resulting in dysregulation of eicosanoid immune mediators

**DOI:** 10.1101/2020.07.09.20149849

**Authors:** Benjamin Schwarz, Lokesh Sharma, Lydia Roberts, Xiaohua Peng, Santos Bermejo, Ian Leighton, Arnau Casanovas Massana, Shelli Farhadian, Albert I. Ko, Yale IMPACT Team, Charles S. Dela Cruz, Catharine M. Bosio

## Abstract

The COVID-19 pandemic has affected more than 10 million people worldwide with mortality exceeding half a million patients. Risk factors associated with severe disease and mortality include advanced age, hypertension, diabetes, and obesity.^1^ Clear mechanistic understanding of how these comorbidities converge to enable severe infection is lacking. Notably each of these risk factors pathologically disrupts the lipidome and this disruption may be a unifying feature of severe COVID-19.^1-7^ Here we provide the first in depth interrogation of lipidomic changes, including structural-lipids as well as the eicosanoids and docosanoids lipid mediators (LMs), that mark COVID-19 disease severity. Our data reveal that progression from moderate to severe disease is marked by a loss of specific immune regulatory LMs and increased pro-inflammatory species. Given the important immune regulatory role of LMs, these data provide mechanistic insight into the immune balance in COVID-19 and potential targets for therapy with currently approved pharmaceuticals.^8^

## Main Text

Lipids function in disease to rearrange cellular signaling structures, modify metabolic processes, absorb reactive species, and act directly as both autocrine and endocrine ligands in the regulation of the immune system. Susceptibility to COVID-19 disease is strongly associated with pre-existing conditions characterized by dysregulation of the lipidome and metabolome.^4-6^ While several studies have examined the systemic metabolic correlates of COVID-19, a well resolved interrogation of the lipidomic changes in COVID-19 severity has not been pursued.^9-12^ To measure lipidomic changes in COVID-19 and gain mechanistic insights into how these changes may drive disease severity, we used serum draws from 19 healthy patients (healthy), 18 COVID-19 patients who did not require ICU admission (moderate) and 20 patients that required ICU admission (severe). The demographics, preexisting conditions, and treatment details of these patients are indicated in Table 1. Lipid and metabolite measurements were made using a series of targeted LC-MS/MS methods providing high-confidence feature identification.^13,14^ Importantly for lipidomic analysis, this enabled the resolution of acyl-chain length and degree of unsaturation, which are both essential for understanding structural and functional rearrangement of the lipidome.

**Table 1.** Patient demographics, preexisting conditions and treatment distributions. Patient data were compared using the chi-square test or Fisher’s exact test for categorical variables and one-way analysis of variance (ANOVA) or unpaired t test was used for continuous variables.

|  | Healthy<br>(n=19) | Moderate<br>(n=18) | Severe<br>(n=20) | p-value |
| --- | --- | --- | --- | --- |
| **Demographics** |  |  |  |  |
| **Mean Age ± SD**<br>**(Range)** | 42.7 ± 14.92<br>(21-72) | 60.56 ± 14.11<br>(26-92) | 63.6 ± 15.17<br>(32-91) | <0.001 |
| **Mean BMI ± SD**<br>**(Range)** | | 29.65 ± 8.37<br>(19.49-55.04) | 31.08 ± 5.84<br>(22.87-44) | 0.5402 |
| **Gender, n (%)** |  |  |  | 0.6391 |
| Male | 9 (47.37%) | 11 (61.11%) | 12 (60%) |  |
| Female | 10 (52.63%) | 7 (38.89%) | 8 (40%) |  |
| **Race, n (%)** |  |  |  | 0.8932 |
| White | 15 (78.95%) | 13 (72.22%) | 14 (70%) |  |
| Black | 2 (10.53%) | 2 (11.11%) | 3 (15%) |  |
| Hispanic | 1 (5.26%) | 2 (11.11%) | 3 (15%) |  |
| Asian | 1 (5.26%) | 1 (5.56%) | 0 (0%) |  |
| **Comorbidities** |  |  |  |  |
| Heart disease |  | 5 (27.78%) | 10 (50%) | 0.1983 |
| Hyperlipidemia |  | 4 (22.22%) | 6 (30%) | 0.719 |
| Hypertension |  | 10 (55.56%) | 15 (75%) | 0.3071 |
| Chronic lung diseases |  | 1 (5.56%) | 4 (20%) | 0.3436 |
| Diabetes |  | 5 (27.78%) | 7 (35%) | 0.7342 |
| **Therapies** |  |  |  |  |
| Tocilizumab |  | 9 (50%) | 18 (90%) | 0.0113 |
| Hydroxychloroquine |  | 13 (72.22%) | 20 (100%) | 0.0171 |
| Steroids |  | 2 (11.11%) | 8 (40%) | 0.0673 |
| **Antiviral** |  |  |  | 0.5292 |
| Azatanavir |  | 11 (61.11%) | 13 (65%) |  |
| Remdesivir trial |  | 2 (11.11%) | 4 (20%) |  |
| **Mechanical ventilation** |  | 0 (0%) | 13 (65%) | <0.0001 |
| **Intensive Care Unit** |  | 0 (0%) | 20 (100%) | <0.0001 |

Changes in primarily polar metabolites among COVID-19 patient cohorts from China, Italy, and France have been reported.^9-12^ In agreement with those studies, we observed a dysregulation of amino acid pools, interruption of the glucose to lactate balance, and dysregulation of nucleotide catabolic products such as xanthine, hypoxanthine, and urate (Sup. Fig. 1a-d).^9-12^ These indicators suggest a robust xanthine oxidase stress response, associated with heart disease,^15-17^ and likely reflect the degree of hypoxia/hypoxemia in the patient, which is a known to be associated with COVID-19 mortality.^18-22^ These data also indicate broad agreement across international populations in metabolic correlates of COVID-19.

We next measured lipidomic profiles across severe and moderate COVID-19 infection along with the healthy controls. To ensure comprehensive recovery of lipid classes, we utilized a modified chloroform extraction method to recover both neutral and polar lipids.^23-25^ By unbiased principle component analysis (PCA), infected patients segregated from healthy in the negative ionization dataset but overlapped in the positive ionization dataset (Sup. Fig. 2a-d). Group-biased partial least square discriminant analysis (PLSDA) of the combined lipid dataset shows non-overlapping healthy and infected separation across the primary axis of variance and a subgroup of severe patients that separate across the secondary axis of variance (Fig. 1a). Specifically, the infected cohorts were associated with increased levels of free poly-unsaturated fatty acids (PUFAs), rearrangement of certain sphingomyelins, and decreased levels of PUFA-containing plasmalogens (Fig. 1b). Parallel univariate analysis revealed that numerous neutral lipids significantly varied between severe and healthy controls, which may reflect either changes in metabolism during infection or pre-infection differences in lipid levels due to pre-existing conditions such as obesity (Sup. Fig 2e-g). Minor patterns distinguishing both infected from healthy cohorts and moderate from severe disease were observed across lysophospholipids (Sup. Fig. 2h-j), cholesterol esters (chol-est) (Sup. Fig. 2k-m), and sphingolipids (Sup. Fig. 2n-p).

**Figure 1.**
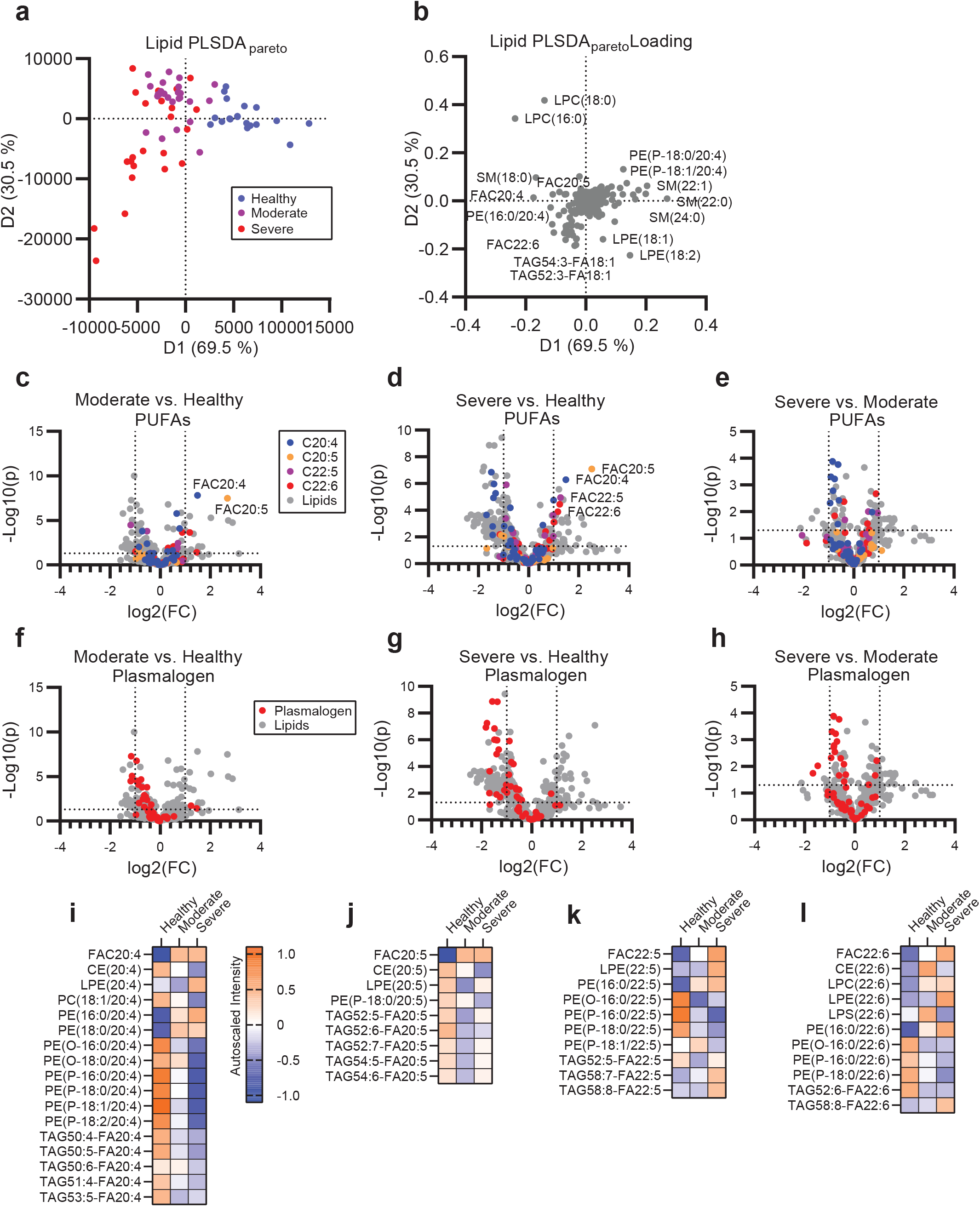
Mobilization of plasmalogen-derived PUFAs correlates with the disease severity in COVID-19. **(a)** Supervised PLSDA analysis of the healthy, moderate and severe disease groups and **(b)** the corresponding feature loading plot. (c-e) Comparison of moderate to healthy **(c)**, severe to healthy **(d)** and severe to moderate **(e)** by unpaired t-test with PUFA classes overlaid. Overlaid data series correspond to lipid species containing at least one copy of C20:4 (blue), C20:5 (orange), C22:5 (purple) or C22:6 (red) acyl chains. **(f-h)** Comparison of moderate to healthy **(f)**, severe to healthy **(g)** and severe to moderate **(h)** by unpaired t-test with plasmalogen lipid series overlaid in red. For (c-h) cutoff lines indicate a positive or negative 2-fold change and a p-value of 0.05. **(i-l)** Heatmap of the autoscaled mean intensity of each patient group for significantly varied lipids (p<0.05) containing C20:4 **(i)**, C20:5 **(j)**, C22:5 **(k)**, and C22:6 **(l)**. Color scale is consistent for (i-l).

Across all lipid classes, PUFA containing lipids were abundant amongst the pool significantly varied lipids between COVID-19 patients and healthy controls (Fig. 1c-e). To examine the regulation of PUFAs between lipid classes and patient groups, we categorized lipids containing C20:4, C20:5, C22:5 or C22:6, which likely represent arachidonic acid (AA), eicosapentaenoic acid (EPA), docosapentaenoic acid (DPA), and docosahexaenoic acid (DHA). Of these PUFA-containing families, changes in the C20:4 series were overrepresented in infected cohorts and could distinguish the severe from the moderate disease patients (Fig. 1c-e). Many of these differentially regulated C20:4 species were the same plasmalogen species that drove separation between infected and healthy cohorts by PCA (Sup. Fig. 2b) and PLSDA (Fig. 2b, f-h). Interestingly, the depletion of PUFA-containing plasmalogen and increased levels of the corresponding free-fatty acids (FAC) indicates the disease progression from the moderate to the severe disease across each PUFA family (Fig. 1i-l). Plasmalogen is known to be a primary pool of PUFAs in both immune and structural cells.^2,26^ Upon systemic immune activation, PUFAs are liberated from their parent glycerolipids and subsequently converted to a wide variety of immune signaling eicosanoids and docosanoids.^27-32^ The balance between pro-inflammatory, immune-regulating, and pro-resolving lipid mediators can drastically change the efficacy of the immune response during infectious and sterile inflammatory diseases as well as during the successful resolution of inflammation following disease.^33-35^ Therefore, we assessed the correlation of the eicosanoid and docosanoid species with COVID-19 disease severity. We targeted 67 eicosanoid and docosanoid species using LC-MS/MS and 15 cytokines by Cytometric Bead Array (CBA) or ELISA to relate lipid changes to markers of disease severity. Eicosanoid and docosanoid lipid mediator (LM) signals were assessed by comparison to standards and available spectral libraries (Sup. Fig. 3-5).^36^ PCA analysis of the combined LM and cytokine data showed separation of infected and healthy cohorts and overlapping, but distinct, separation between moderate and severe patients (Fig. 2a). Nearly all LMs measured were positively correlated with infection (Fig. 2b). Univariate analysis showed significant enrichment of the majority of LMs measured in both the moderate and severe groups (Fig. 2c, d). Interestingly, moderate and severe disease were characterized by unique milieus of LMs and cytokines (Fig. 2b, e). Moderate disease was characterized by significantly higher levels of the pro-resolving LM resolvin E3 (RvE3). Further, there was a trend toward increased presence of the prostaglandin family members, particularly PGE2 (p= 0.105), PGFD2 (p= 0.220) and PGF2a (p= 0.242). In contrast, severe disease was characterized by a further increase in free PUFAs levels, AA-, EPA-, DPA and DHA-derived mono-hydroxylated species and AA-derived dihydroxylated species (Fig. 2c-e). This shift in specific immune regulatory LMs in severe disease suggests that an imbalance of LMs may contribute to disease progression. LMs are generated by a single or a series of oxygenase mediated conversions of the parent PUFA. To examine the potential contribution of each oxygenase enzyme to the severe disease phenotype we grouped LMs according to synthesis pathway (Fig. 2g-k). Several LMs are shared between multiple enzyme groups as they require sequential stereospecific hydroxylations. This grouping revealed that moderate disease was characterized by higher cyclooxygenase activity (COX) as well as certain products of ALOX12 while severe disease is characterized by greater activity of ALOX5 and cytochrome p450 (CYP) enzymes. This is good agreement with previous observations from influenza, which associated symptom severity with ALOX5 activity.^37^

**Figure 2:**
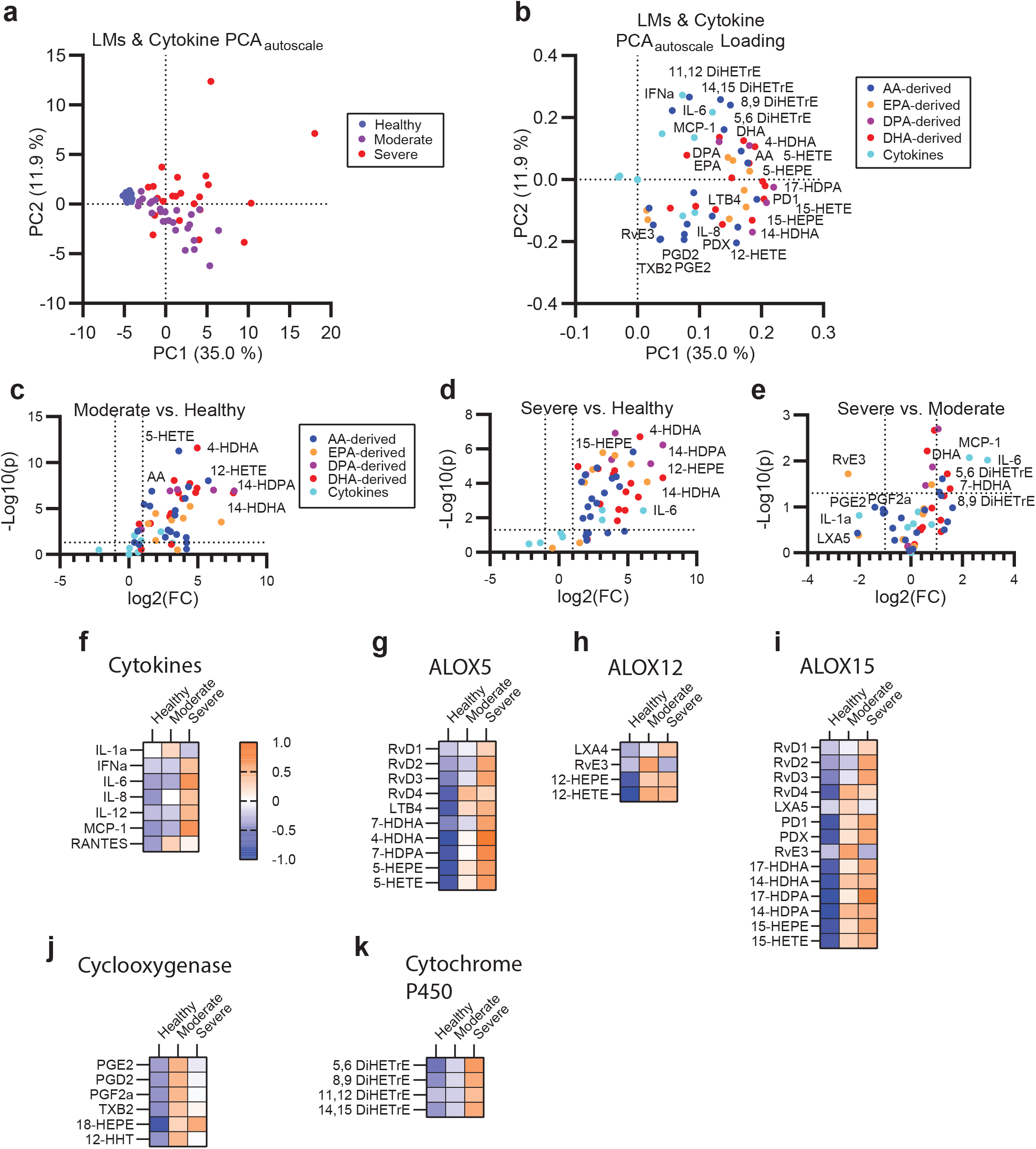
A unique milieu of LMs defines moderate and severe COVID-19 disease. **(a)** Unsupervised PCA of autoscaled combined lipid mediator and cytokine dataset and **(b)** corresponding feature loading plot. **(c-e)** Univariate comparison of moderate disease to healthy **(c)**, severe disease to healthy **(d)** and severe to moderate disease **(f)** by unpaired t-test. Cutoff lines indicate a positive or negative 2-fold change and a p-value of 0.05. For (b-f) species are colored by class as cytokine (cyan), arachidonic acid-derived (AA-derived, blue), eicosapentaenoic acid-derived (EPA-derived, orange), docosapentaenoic acid-derived (DPA-derived, purple) or docosahexaenoic acid-derived (DHA-derived, red). **(f-k)** Heatmaps of the autoscaled mean for each patient group across cytokines **(f)**, molecules synthesized by ALOX5 **(g)**, ALOX12 **(h)**, ALOX15 **(i)**, Cyclooxygenases **(j)** or cytochrome P450 **(k)**. Color scale is consistent across (f-k).

COVID-19 comorbidities including obesity, age, heart disease, and diabetes are characterized by dysregulation of the homeostatic lipidome.^1,3^ To assess the correlation of these conditions with the shifts in LM pools and their glycerolipid precursors, we overlaid age, sex, BMI, diabetes, heart disease, survival, and antiviral treatment onto the separate PCAs (Sup. Fig. 7-15). Age and sex were evenly distributed across the infected cohort on all PCAs (Sup. Fig. 7, 8). Of the treatments examined, only remdesivir showed a negative correlation with the disease severity (Sup. Fig. 9-11). BMI, diabetes, heart disease and morbidity segregated with the severity of disease across all datasets (Sup. Fig. 12-15). From this study, it is likely that the lipidomic imbalance associated with severe disease is at least partially a consequence of homeostatic disruption of the lipidome due to these pre-existing conditions. It is likely that these pre-existing lipidomic imbalances are further exacerbated during COVID-19 through dysregulation of the LM response resulting in severe disease, impaired resolution and persistent inflammation.

Elevation of ALOX5- and CYP-dependent LMs in severe COVID-19 patient sera suggested systemic upregulation of these pathways. To examine the cellular origin of these enzymes in COVID-19 patients, we interrogated a published single cell RNAseq dataset of COVID-19 patient PBMCs for expression of ALOX and CYP genes (Sup. Fig. 6a, b).^38^ *ALOX5* expression was detected in most of the 20 cell types identified (Fig. 3a-b) with the highest expression in CD14 monocytes, CD16 monocytes, neutrophils, B cells, and DCs (Fig. 3c). *ALOX5* expression was significantly increased in neutrophils and trended upward in CD14 monocytes, CD16 monocytes, and developing neutrophils (a population found almost exclusively in diseased individuals) from COVID-19 patients compared to healthy controls. Interestingly, severe COVID-19 is characterized by elevated *ALOX5* expressing monocyte/macrophage population and depletion of lymphocyte populations.^38-40^ The absence of CYP genes in the blood was consistent with the primarily hepatic localization of these enzymes.^41^ These data suggest a systemic dysregulation of ALOX5 and further support the metabolic dysregulation of the liver in severe disease.^42^

**Figure 3.**
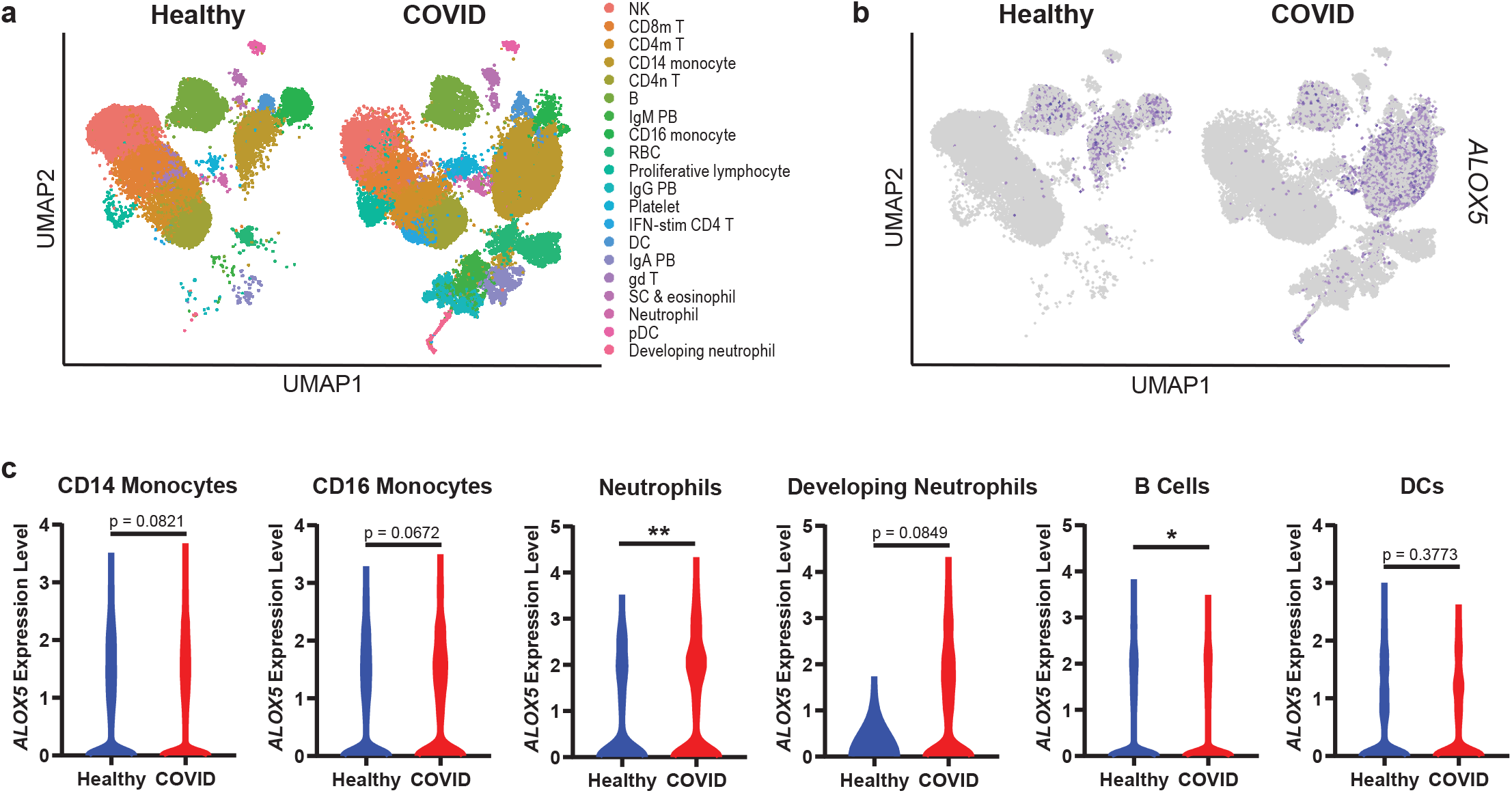
Human PBMCs from COVID-19 patients are enriched for *ALOX5* expressing cells and express higher levels of *ALOX5*. **(a)** UMAP dimensionality reduction plot of a published human PBMC single-cell RNA Seq dataset (Wilk, et al *Nature Medicine* 2020) identifying twenty cell types. **(b)** UMAP depicting *ALOX5* expressing cells in blue. **(c)** Violin plots indicating *ALOX5* expression levels within specific cellular populations in healthy (blue) or COVID (red) PBMCs. Statistical significance was determined by a Mann-Whitney test; * p < 0.05. ** p< 0.01.

## Conclusion

These results provide the first detailed lipidomic understanding of COVID-19 disease progression and represent one of the first combinations of bulk lipidomic and eicosanoid data to map mobilization of lipids in human infectious disease.^7,37^ We provide evidence that a systemic lipid network consisting of liberated PUFAs from plasmalogen and their subsequent conversion to LMs, capable of modulating inflammatory responses, characterizes both the onset and severity of COVID-19. Specifically, the loss of the immune regulatory prostaglandins and the increased production of AA-derived products of ALOX5 and cytochrome P450 provides both a measure of disease severity and a mechanistic understanding of the immune balance allowing for patient recovery. ^43^ Importantly, these pathways are directly targetable with drugs previously approved for use in other inflammatory conditions and, thus, provide therapeutic opportunities to control severe COVID-19.^27,31^

## Data Availability

All data is posted and/or available upon request

## Abbreviations

LM: eicosanoid and docosanoid lipid mediators
PE: phosphatidylethanolamine
LPE: lyso-PE
PC: phosphatidylcholine
LPC: lyso-PC
PS: phosphatidylserine
PE(O) or PE(P): plasmenyl or plasmanyl plasmalogen
TAG: triacylglycerol
DAG: diacylglycerol
MAG: monoacylglycerol
CE: cholesterol ester
Cer: ceramide
DCer: dihydroceramide
HCer: hexosylceramide
LCer: lactosylceramide
SM: sphingomyelin
FAC: frees fatty acid
Rv: resolvin
LX: lipoxin
LT: luekotriene
HETE: hydroxyeicosatetraenoic acid
HEPE: hydroxyeicosapentaenoic acid
HDHA: hydroxydocosahexaenoic acid
HDPA: hydroxydocosapentaenoic acid
PG: prostaglandin
PD: D-series protectin
TxB2: Thromboxane B2
LC-MS/MS: liquid chromatography tandem mass spectrometry
CBA: cytometric bead array
PCA: principle component analysis
PLSDA: partial least square discriminant analysis

## Acknowledgements

We are deeply indebted to the patients and families of patients for their contribution to this study. Prof. Charles N. Serhan and his group including, K. Boyle, A. Shay, C. Jouvene, X. de la Rosa, S. Libreros, and N. Chiang generously provided methodology, consultation and extensive training for the assessment of lipid mediators by LC-MS/MS. AB Sciex in particular M. Pearson, P. Norris and P. Baker (currently Avanti Polar Lipids) provided LC-MS/MS consultation and methods. Lokesh Sharma is supported by Parker B Francis Fellowship. Charles Dela Cruz is supported by Veterans affairs Merit Grant (BX004661) and Department of Defense grant (PR181442). Albert Ko and Charles Dela Cruz are supported by a U19 supplement for this work (AI089992-09S2). This work was supported by the Intramural Research Program of the National Institutes of Health, National Institute of Allergy and Infectious Diseases. This work was supported by the Department of Internal Medicine at the Yale School of Medicine, Yale School of Public Health, and the Beatrice Kleinberg Neuwirth Fund.

## Author Contributions

B.S., L.S., C.D.C and C.M.B. conceived of experiment. L.S., X.P., S.B., A.C.M, S. F. and A.I.K and the Yale IMPACT Team enrolled patients and collected samples. B.S., L.R. and I.L. extracted samples and collected data. B.S. conducted metabolomics and lipidomics analysis. L.R. conducted single cell RNAseq analysis and cytokine analysis. B.S., L.R., L.S., C.D.C. and C.M.B wrote the manuscript. Yale Impact team: (Listed in alphabetical order) Kelly Anastasio, Michael H. Askenase, Maria Batsu,, Sean Bickerton, Kristina Brower, Molly L. Bucklin, Staci Cahill,, Yiyun Cao, Edward Courchaine,, Giuseppe DeIuliis, John Fournier, Bertie Geng, Laura Glick, Akiko Iwasaki, Nathan Grubaugh, Chaney Kalinich, William Khoury-Hanold, Daniel Kim, Lynda Knaggs, Maxine Kuang, Eriko Kudo, Joseph Lim, Melissa Linehan, Alice Lu-Culligan,, Anjelica Martin, Irene Matos, David McDonald, Maksym Minasyan, M. Catherine Muenker, Nida Naushad, Allison Nelson, Jessica Nouws,, Abeer Obaid, Camilla Odio, Saad Omer, Isabel Ott, Annsea Park, Hong-Jai Park, Mary Petrone, Sarah Prophet, Harold Rahming, Tyler Rice, Kadi-Ann Rose, Lorenzo Sewanan, Denise Shepard, Erin Silva, Michael Simonov, Mikhail Smolgovsky,, Nicole Sonnert, Yvette Strong, Codruta Todeasa, Jordan Valdez, Sofia Velazquez, Pavithra Vijayakumar, Annie Watkins, Elizabeth B. White, Yexin Yang

## Materials and Methods

### Ethics Statement

This study was approved by Yale Human Research Protection Program Institutional Review Boards (FWA00002571, Protocol ID. 2000027690). Informed consents were obtained from all enrolled patients. The healthy blood samples were obtained under the protocol (HIC 0901004619) before the onset of COVID-19 outbreak.

### Chemicals

Tributylamine was purchased from Millipore Sigma. LCMS grade water, methanol, isopropanol, chloroform and acetic acid were purchased through Fisher Scientific. All lipid mediator standards were purchased from Cayman Chemical.

### Kits and Reagents

CBA kits were purchased from BD Biosciences.

### Patient cohort and serum collection

Patients were recruited among those who were admitted to the Yale-New Haven Hospital between March 18th and May 9th, 2020 and were positive for SARS-CoV-2 by RT-PCR from nasopharyngeal and/or oropharyngeal swabs. Patients in this study were enrolled through the IMPACT biorepository study after obtaining informed consent. Basic demographics and clinical information of study participants were obtained and shown in Table 1.

Prior to thawing, all samples were gamma-irradiated (2 MRad) to inactivate potential infectious virus.

### Sample processing for aqueous, organic, and lipid mediator extraction

For all LCMS methods LCMS grade solvents were used. Sample order was randomized throughout each extraction. For aqueous and organic metabolites, 50 µL patient serum was aliquoted directly into 400 µL of ice-cold methanol and 500 µL of ice-cold chloroform was added. Samples were agitated by shaking for 20 minutes at 4 °C and subsequently centrifuged at 16k xg for 20 minutes at 4 °C to induce layering. The top (aqueous) layer and bottom (organic layer) were collected. The aqueous layer was diluted 1:10 in 50% methanol in water and prepared for LCMS injection. The organic layer was taken to dryness in a Savant™ DNA120 SpeedVac™ concentrator (Thermo Fisher) and stored at −80 °C until analysis. At time of analysis, samples were resuspended in 500 µL of 5 µg/mL butylated hydroxytoluene in 6:1 isopropanol:methanol and further diluted 1:3 in the same solvent combination for LC-M/MS injection.

### Lipid mediators sample processing and extraction

Lipid mediators were extracted from patient serum as previously described.^44^ Briefly 100 µL of serum was aliquoted on ice and 1 ng each of d8-5-HETE, d5-RvD2, d5-LXA4, d4-LTB4, d4-PGE2 was added to each sample followed by 400 µL of ice-cold methanol. Samples were incubated for 30 min at −20 °C to allow precipitation of protein. Samples were centrifuged at 10k xg for 10 minutes and the supernatant was collected in a fresh tube.

Solid phase extraction columns (Sep-Pak^®^ 3 mL, 200 mg, C18, Waters Corporation) were conditioned in vacuum manifold with 10 mL of methanol followed by 10 mL of water. One at a time to each supernatant, 9 mL of acidified water (pH 3.5 with hydrochloric acid) was added and the samples was quickly loaded onto column. The column was then washed to with 10 mL of water. Once samples were loaded, columns were washed with 4 mL of hexanes and then lipid mediators were eluted with 8 mL of methyl-formate. Samples were dried under nitrogen and resuspended in 100 µL of 1:1 water:methanol. For LC-MS analysis 30 µL of each sample was injected.

### LC-MS/MS analysis

Aqueous metabolite, lipid, and lipid mediator samples were analyzed using a series of targeted multiple-reaction monitoring (MRM) methods. All samples were separated using a Sciex ExionLC™ AC system and analyzed using a Sciex 5500 QTRAP^®^ mass spectrometer.

Aqueous metabolites were analyzed using a previously established ion pairing method with modification.^14^ Quality control samples were injected after every 10 injections and assessed for signal stability. Samples were separated across a Waters Atlantis T3 column (100Å, 3 µm, 3 mm X 100 mm) and eluted using a binary gradient from 5 mM tributylamine, 5 mM acetic acid in 2% isopropanol, 5% methanol, 93% water (v/v) to 100% isopropanol over 15 minutes. Analytes were detected in negative mode using two distinct MRM pairs for each metabolite when possible. After signal confirmation only one of the MRM signals was taken forward for analysis. Heavy labeled standards were not utilized given the breadth of targets, thus relative quantification was performed. Fidelity of select signals including retention time and spectra was confirmed by comparison to a synthetic molecular reference.

Lipid samples were analyzed using a previously established HILIC method with modification.^13^ Samples were separated on a Water XBridge^®^ Amide column (3.5 µm, 3 mm X 100 mm) and eluted using a 12 minute binary gradient from 100% 5 mM ammonium acetate, 5% water in acetonitrile apparent pH 8.4 to 95% 5 mM ammonium acetate, 50% water in acetonitrile apparent pH 8.0. Target lipids were detected using scheduled MRM. Lipid signals were divided into two methods utilizing either negative mode or positive mode and a separate injection was analyzed for each method. Both datasets were separately normalized using total-area sum to correct for instrument drift.

Lipid mediators were analyzed using a previously established reverse phase method with modifications.^44^ Samples were separated on a Waters Atlantis T3 column (100Å, 3 µm, 3 mm X 100 mm) and using a binary gradient of A: 0.01 % acetic acid in water and B: 0.01 % acetic acid in methanol. Samples were eluted over 20 min from 40-100 % B. Samples were detected in negative mode using previously published MRM pairs and source conditions.^36^ Triggered spectra were collected using enhanced-product ion scans and rolling collision energy. A blank and a standard mix were serially injected every 10 injections. Standard mix consisted of each of the following compounds at 10 ng/mL: RvE3, LXA4, LXA5, LXB4, PGE2, PGD2, PGF2a, TxB2, PD1, RvD5, Maresin 1, LTB4, 5,15-DiHETE, 14-HDHA, 18-HEPE, AA, EPA, DHA. Spectra and comparison to authentic standards was used to confirm signal identity.

Spectral confirmation was not possible for RvD2, RvD3, LXA5, RvD6, 8,9 DiHETrE, 12-HHT, 11-HETE, 11-HEPE, 7-HDPA, 13-HDPA, 14-HDPA, 17-HDPA, 7-HDHA, 13-HDHA, 17-HDHA and 21-HDHA but identity was assessed by comparison to related standards. These signals were regarded as lower confidence but were used for class comparison of the LMs and multivariate analysis.

All signals were integrated using MultiQuant^®^ Software 3.0.3. In total 1,414 molecules were targeted across a water-soluble metabolite method and two organic-extracted lipid methods in either positive or negative ionization modes. Of these 716 features were judged to be positively detected by visual inspection, missing value filtering (50% cut-off) and QC coefficient of variance filtering (40% cut-off after normalization). Remaining missing values were replaced with the minimum group value for that feature. For aqueous and lipid mediator datasets signal quality was judged visually and signal stability was assessed by QC or repeat injection of a standard mix. Lipid mediator data was normalized to internal heavy isotope standards as previously described.^44^

Univariate and multivariate analysis was performed in MarkerView^®^ Software 1.3.1. The aqueous dataset and the combined lipid mediators/cytokine dataset data were autoscaled prior to multivariate analysis in order to visualize the contribution of low ionization efficiency species and difference of scales between the cytokine and lipid mediator measurements. Lipid datasets were pareto scaled to avoid overrepresenting low abundance signals within each lipid class. For all univariate analysis an unpaired t-test was used. For univariate analysis of the LM/cytokine set a single moderate group patient was excluded by an extreme studentized deviate test for analysis of PGE2 (z = 4.58).

### Quantification of cytokine and chemokine levels

The serum concentration of IFN-α, IFN-γ, IL-1β, IL-2, IL-4, IL-6, IL-8, IL-10, IL-12/IL-23p40, IL-17A, MIP-1α, RANTES, TNF-α, and MCP-1 was determined using a cytometric bead array according to the manufacturer’s instructions (BD Biosciences). The serum concentration of IL-1α was determined by an ELISA according to the manufacturer’s instructions (R & D Systems).

### Single cell RNA sequencing analysis

The published single cell RNA sequencing dataset from Wilk, et al Nature Medicine 2020 was downloaded from the COVID-19 Cell Atlas (https://www.covid19cellatlas.org/#wilk20).^38^ Data was read into Seurat v3.0 and each cluster’s cellular identity was annotated per Wilk, et al Nature Medicine 2020.^38^ Expression levels of ALOX and CYP genes within specific cell types in healthy controls and COVID-19 patients was visualized using Seurat’s DotPlot feature. *ALOX5* expression levels in specific cell types was visualized using the VlnPlot feature A Mann-Whitney test was used to determine statistical differences in gene expression between healthy and COVID samples.

### Patient Statistics

Demographic data is presented as either counts and percentages (for categorical data) or means and standard deviations (for continuous data). To investigate the difference in the control, moderate and severe groups, GraphPad Prism (version 8.4.2) was used. The results were compared using the chi-square test or Fisher’s exact test for categorical variables and one-way analysis of variance (ANOVA) or unpaired t test was used for continuous variables. A p-value of less than 0.05 was considered statistically significant.

